# Dermatophytic Diseases: A Review of Tinea Pedis

**DOI:** 10.1101/2021.06.28.21259664

**Authors:** Gyamfi Agyemang Julien, Wezena Cletus Adiyaga, Rahmat Alela-Emoma Saaka, Samuel Sunwiale Sunyazi, Abraham Titigah Batuiamu, Daniel Abugri, James Abugri

## Abstract

Studies in skin diseases, particularly *Tinea pedis* are very rare in Ghana leading to low investment in dermatological services in the country and the African continent. Globally, Tinea pedis affects about 15% of the world’s population. Importantly, it is a major public health problem and socioeconomic issue. Currently, the most recommended treatment for *Tinea pedis infection are* polyenes, azoles, allylamines, and fluorocytosines. Although these drugs are effective, they do have adverse side effects and are limited in the clinical settings in developing countries especially Ghana.

**Method:** Research papers were collected from Pubmed, Google scholar, chemical abstracts, and journal websites, reporting both *in vitro* and *in vivo* information on Tinea pedis. General information on Tinea pedis, the methods of infection, transfer, treatment options, and resistance were obtained after screening the articles. Many agents are involved in cases of Tinea pedis but are predominantly caused by *Trichophyton rubrum* which feeds on the keratinous layer of the topmost skin of the foot causing skin discoloration, itching, and maceration. The disease is influenced by individual lifestyle, environmental conditions, and individual factors such as age, sex, and host immunity and is prevented by the maintenance of good personal hygiene.

Tinea pedis like other tinea infections are treated with both topical and systemic agents. The classes of medications used in the treatment of Tinea pedis are polyenes, azoles, allylamines, and fluorocytosines. Tinea pedis antifungal resistance development could be partly associated with incomplete medication and misuse of antifungal medications. Additionally, patients with serious underlying health conditions such as compromised immune systems like HIV/AIDS, diabetes, radiotherapy for cancer, and transplantation could complicate resistance.

**Conclusion:** Although, fungal diseases do not cause epidemics the increasing rate of fungal infections and therefore Tinea pedis has to be checked and prevented. High budgets are made in the development of medications which mostly lose their effectiveness over time due to resistance development. Good personal hygiene is very effective but the available medications must be used appropriately for effective treatment and resistance avoidance. Infection prevention and control, tracking and data sharing, good and easily accessible antifungals, vaccines, and maintenance of personal and environmental hygiene are the topmost preventive measures against resistance development.

## Introduction

Globally, it has been estimated that 15% of the world’s human population are infected with fungal infections^1,2^ with more than 70% having a high chance of getting infected by a fungal infection over their lifetime^1^. Fungal infections affect not only humans but also animals, plants, and cause havoc to the ecosystem^3^. Dermatophytes are a class of fungi that require keratin for growth and thus infect and colonize keratinous layers of the skin, nails, and hairs^4,5,6^. Tinea pedis or athlete’s foot, a common type of dermatophytosis is associated with the skin between the toes but is capable of spreading to other parts of the foot; the sole, sides, and dorsum^7^ and the body; groins, fingernails, and fingers. Tinea means fungal infection whereas pedis is a Latin word for infection of the foot^7^. Other forms of *Tinea* infections include tinea capitis which is an infection of the scalp^8^ also called ringworm of the scalp, tinea corporis which is a contagious infection of the body, also called ringworm of the body^9^, tinea barbae which is an infection of the beard and mustache area^10^, tinea cruris or Jock itch which is an infection of moist and warm areas of the body including the groin and inner thighs^11^ and tinea unguium also called onychomycosis, which is a dermatophyte infection of the toenails or fingernails^12^. Tinea pedis is very common among the homeless^13^. The disease co-morbid with other infections and thus often under-diagnosed. Due to its coexisting ability, the disease is found in some cases of onychomycosis, and tinea unguium serving as the reservoir of other dermatophytes^14^. When left untreated, the disease could lead to other conditions such as wounds, cellulitis, pain, and difficulty of ambulation^15^. Though the disease responds to inexpensive topical agents^7^, it develops a secondary bacterial infection due to the failure of patients to seek medical attention resulting from their uninformed knowledge of fungal infection^15^. Several drugs and creams are available for clearing *tinea* infections; nonetheless, good personal hygiene is one best way to prevent the disease. Mostly, topical agents are for the treatment of Tinea pedis but systemic agents can treat large area infections. Environmental considerations must be taken into account in selecting the right medication^16^, and for the avoidance of recurrence, treatment must continue for at least a week after the clinical clearing.

### Methodology

The work was done through a review of various publications obtained through online database search engines using PubMed, google scholars, mycology journals, infectious diseases websites, bibliography reviews, and newspaper publications. A total of 506 articles were obtained through the online database originating from several countries across the globe with the publication dates ranging from 1971 to 2020. Publications on dermatophytes were minimal in the first 10 years but significantly increased in the last 10 years. Most of the retrieved papers were published starting from the year 2001 to 2020. Publications on dermatophytes were minimal in the first 10 years but significantly increased in the last 10 years. Most of the retrieved papers were published between the years 2001 and 2020. The search topics were Tinea pedis, antifungal susceptibility of Tinea pedis, dermatophytes, dermatophytes infections, and the prevalence of Tinea pedis in Ghana, Africa, and the world. The articles included works from other dermatophyte infections, bacterial, viral, and non-dermatophyte infections in addition to the research topic, Tinea pedis.

**Figure 1:**
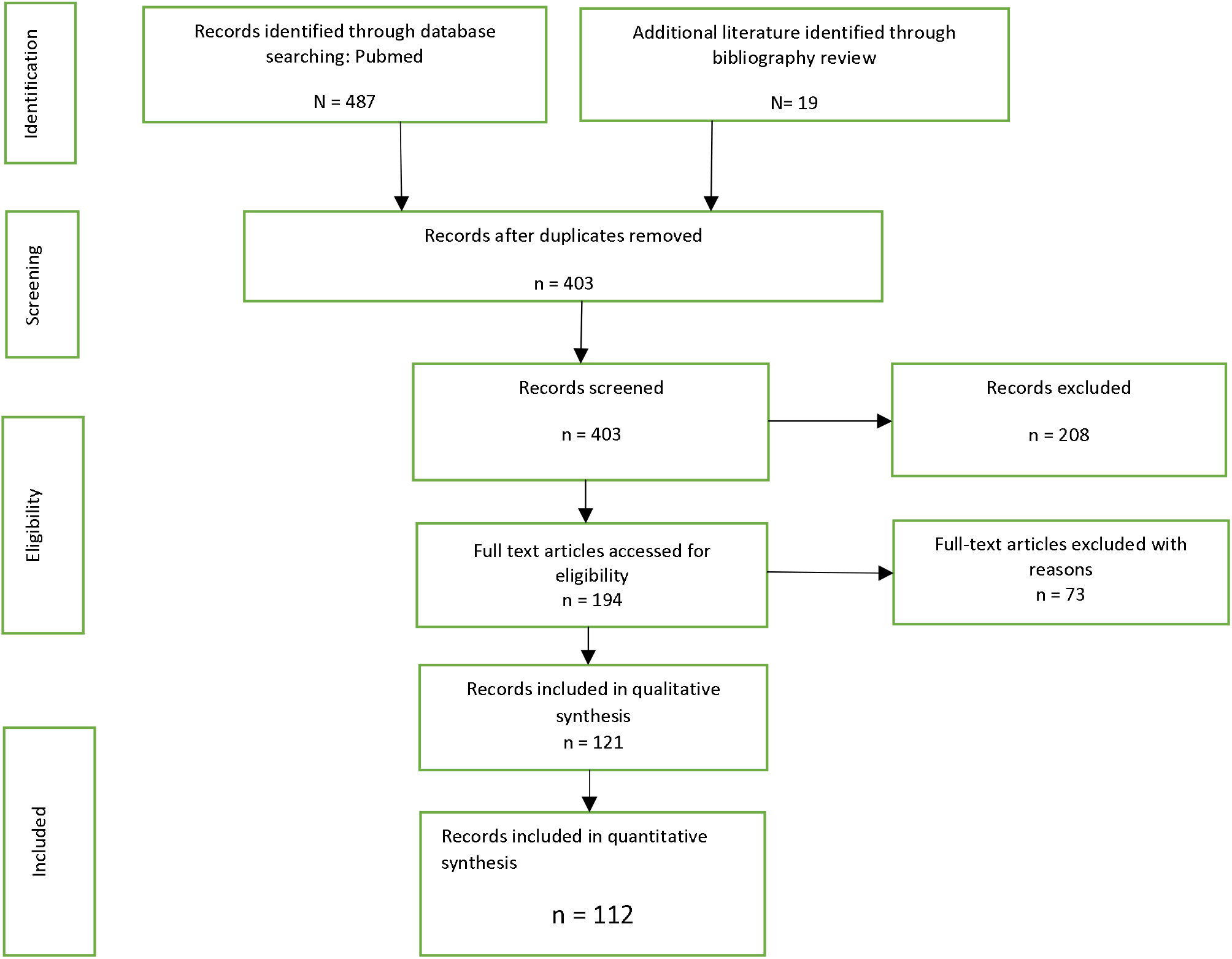
A PRISMA flow diagram of the screening procedure employed in identifying the various articles used in this review.

**Figure 2:**
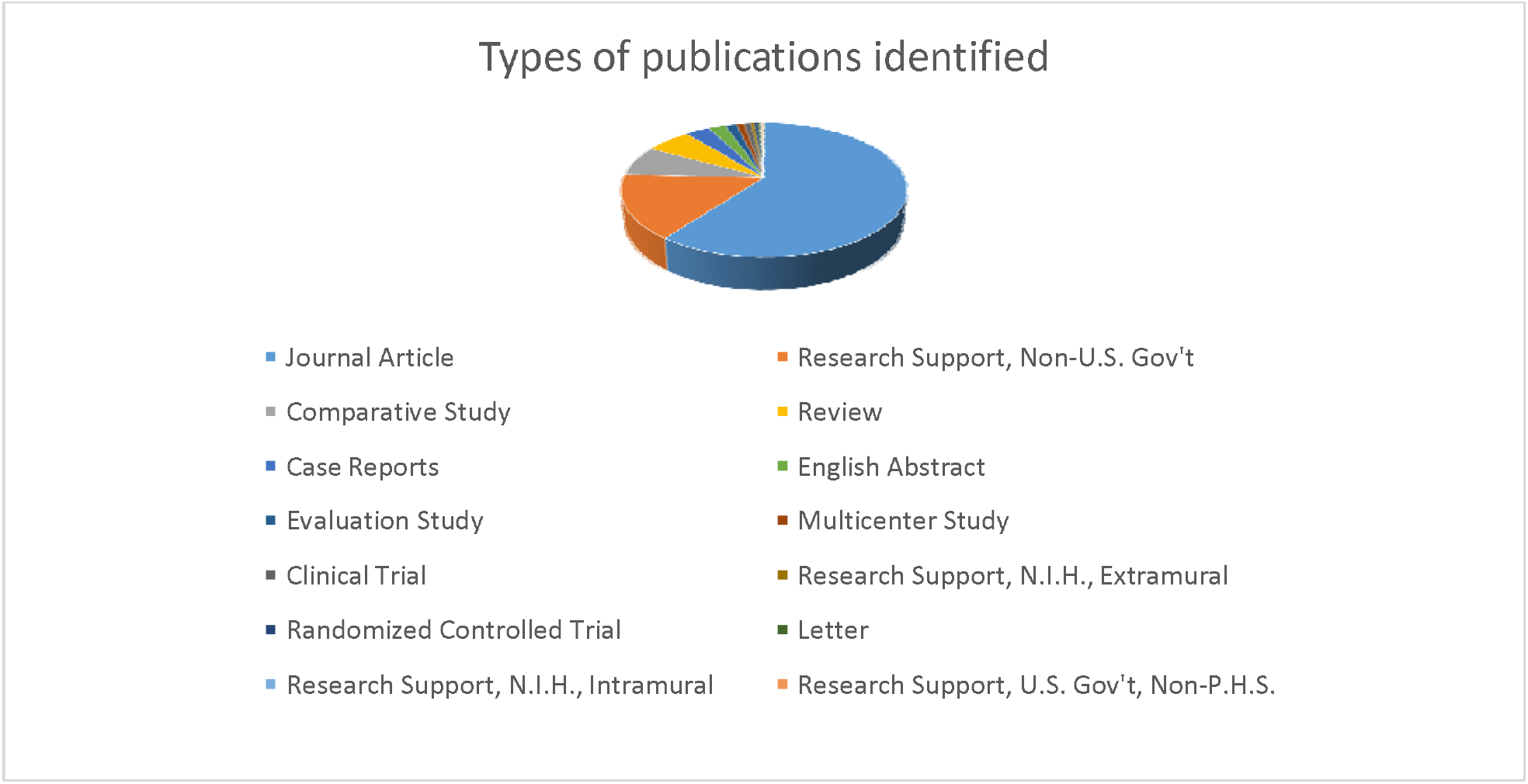
Publication types identified for the work

**Figure 3:**
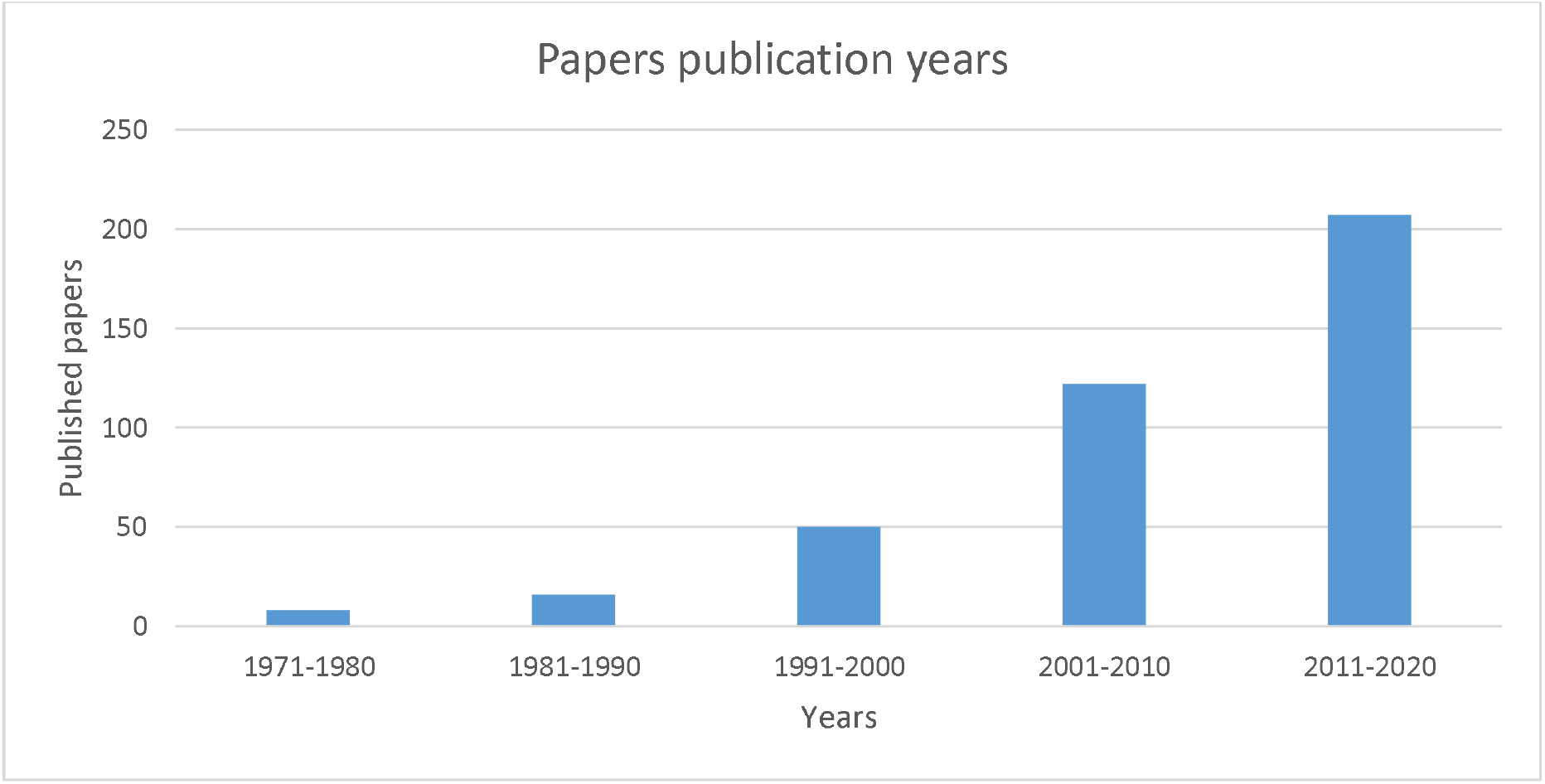
Grouped records of the years of publications of the publications used.

## Classification of causative agents

Dermatophytes are the main agents in Tinea pedis infection^1,7^. They cause skin and nail infection by infecting keratin of the top layer of the epidermis^1^. Fungi, the kingdom under which dermatophytes are classified, exist in the form of unicellular yeast and mold with highly branching structures called hyphae^13^. Dermatophytes are classified as antropophilic, which are those that cause infection only in humans and are transmitted from human to human, zoophilic, which includes those that infected animals but can cause inflammatory responses in humans who come in contact with infected animals, or geophilic, which are mainly found in the soil but occasionally cause infection in humans and animals^17,18^. They are grouped into three asexually reproducing forms; Epidermophyton, Microsporum and Trichophyton based on their primary location of interaction^16,19^. Among these genera, the Trichophyton contain the species that are most implicated in human dermatophytosis^20^. The main causative agent of interdigital Tinea pedis is the antropophilic species, Trichophyton rubrum^17,18^ with other agents such as *T. mentagrophytes*^21^, *Epidermophyton floccosum*, and *T. tonsurans* which rarely occur in adult cases^4^. *T. rubrum* is found in some cases of onychomychosis^22^.

### Skin Microbiome Characterization

Normal foot skin is colonized by harmless commensal microorganisms that include bacteria and fungi^23^. These microorganisms thrive very well in the distinct ecological niche of the foot that has a large number of sweat glands but lacks sebaceous glands that secrete antimicrobial fluids^23^. The warm and moist foot environment fostered by wearing closed shoes promotes the growth of fungi and gram-negative bacteria^23^. Xiaoping Liu and colleagues identified *Corynebacterium tuberculostearicum* and *Staphylococcus pettenkoferi, Pseudoclvibacter alba*, and *Brevibacterium paucivorans* as the prevalent bacteria species found on the feet of healthy individuals^23^. Comparatively, *Corynebacterium tuberculostearicum, Staphylococcus pettenkoferi*, C. minutissimum, and *P. sphaerophysae* were prevalent in patients with tinea pedis. *Cladosporium halotolerans, Phoma saxea, Aspergillus cibarius*, and *Rhodotorula mucilaginosa* are the most prevalent fungal species isolated from the feet of healthy individuals. In patients of tinea pedis, *Trichophyton rubrum, Cladosporium halotolerans, Candida parapsilosis*, and *Wallemia sebi* were the most abundant species isolated^23^. Tinea pedis presents a diverse community of microbes in different patients and different clinical presentations^23^ as reported by Liu et. al^24^. The disease, as shown above, is associated with and complicated by the presence of other infecting organisms^23^. The diversity of bacteria and fungi species was shown to be higher in healthy controls than in tinea pedis patients, even though Staphylococcus and Trichophyton species were the most abundant bacteria and fungi species respectively^23^ The most prevalent strain of fungi in cases of tinea pedis is *Trichophyton rubrum* (~60% of cases)^25^ followed by *T. mentagrophytes*^26^ (~20-30%), *Epidermophyton floccosum* (~10%)^1,6,16,27^ and *T. tonsurans* which rarely occurs in adult cases.

## Clinical presentation

There are three clinical forms of Tinea pedis; interdigital, hyperkeratotic, and vesticular, which are classified based on their pattern of skin manifestations^2,13,16,28^. Vikas and colleagues^1^, however, classified tinea pedis into four clinical forms with the fourth class being ulcerative tinea pedis.

Interdigital Tinea pedis is the most common type of dermatophytes and occurs between the toes, mainly the fourth and fifth toes. It presents as macerated, dry or wet, scaly, and peeling of the interspaces of the toes and is classified as simplex if dry and asymptomatic, or complex if wet and symptomatic as a result of secondary bacterial infection.

Plantar (Hyperkeratotic) Tinea pedis affects the soles, heels, and sides of the foot with characteristic powdery scaling, redness of the soles, heels, and sides of the foot. The involvement of the plantar and lateral aspects of the foot with erythema and hyperkeratosis characterizes the moccasin foot.

Vesicobullous Tinea pedis occurs on the soles of the foot. It presents with an acute inflammatory reaction consisting of vesicles, pustules^29^, or blisters.

A patient may be asymptomatic or symptomatic by experiencing burning or itching. Infected toenails appear thickened, discolored, and dystrophic^13^.

Ulcerative tinea pedis is characterized by rapidly spreading ulcers, erosions, and lesions which typically occur in the web of the toes and occasionally affecting the soles of the foot. Large areas of the foot can be sloughed off. This type of tinea pedis is commonly followed by secondary bacterial infection and is mostly a disease of the immunocompromised and diabetic patients^1^. Symptoms such as malaise, pharyngitis, pyrexia, and cellulitis may accompany the infection.

### Modes of Transmission

Fungi grow well in moist, dark, and warm environments. Poor hygiene, wearing dirty^30^, infected^31^, and occlusive footwear ^1,16,32^, dirty socks, and wet feet provide the right conditions for infection and transmission of tinea pedis^13,33^. Tinea pedis is contagious and is transmitted through contact with fungal spores, arthroconidia from an infected person, surfaces, or objects^1,13^. Pets are a possible source of infection transmission.

### Infection and survival

Infection is mounted through the interaction of the dermatophyte with the keratin of the top layer of the skin^13,19^. The arthroconidia of the dermatophyte invade the resistance mechanism (keratin layer, pH, and mucosal surfaces) of the host and penetrate its hyphae into the keratin layer of the skin. The fungi can adhere to the host due to the mannans in the glycoproteins of their cell wall. On infection, the dermatophytes use the carbons, nitrogen, and sulfur from keratin as energy sources thereby degrading the keratinous layer by the injection of permeases, enzymes of the cell wall, and hydrolytic enzymes. The tinea fungi do not penetrate the living tissues of the human host but colonize the horny keratinous layer^2,19^.

### Prevalence

Fungal infections, in general, have been reported to be on a rapid rise recently even though these infections do not normally cause an outbreak^19^. Fungal diseases affect an estimated 15% of the population at any point in time with approximately 70% getting fungal infections over their lifetime^34^. Tinea pedis is found in almost all regions of the globe^1,13^ and affects all classes of people^2^ if appropriate personal hygiene instructions are not followed. The disease is influenced by individual lifestyle, environmental conditions, and individual factors such as age, sex, and host immunity^14^. An Individual’s lifestyle, which does not obey hygienic procedures, can lead to infection while environmental and climatic conditions influence the condition due to its higher observation in areas with a humid climate, which tends to promote the growth of the pathogen^1^ and lower in places where people do not usually wear shoes^1^. The prevalence of the disease varies with age with adults are being much more susceptible than children^1,2,13,14,35^. The area’s most commonly affected are interdigital or pompholyx^36^ areas of the foot. Also, males are more susceptible to the disease than females of the same or similar age^2,16^. Tinea pedis is also found to be prevalent among army personnel^1^, school children^1,37,38,39,40,41,40,42^, sailors, coal-miners, athletes, those attending public swimming baths^1,37^, or leisure centers^37^.

Dermatophyte infections are severe in people living with Acquired Immune Deficiency Syndrome (AIDS)^1,13,19,37^, diabetes mellitus patients (who have a 50% chance of infection)^1,13,37,43,44^, and those with a suppressed immune system due to medical procedures such as organ transplantation and chemotherapy^1,13,19^. Obesity also influences the severity of Tinea pedis^1^ with non-completion of medication leading to disease recurrence and the development of resistance.

### Diagnosis

Diagnosis of tinea pedis is based on clinical evaluation, laboratory evaluation, or fungal culture. The history of occurrence which would include the review of the patient for exposure to risk factors of the disease and clinical examination for the obvious appearance of the disease areas (symptoms) are important aspects of clinical diagnosis. In instances where appearance will not aid an obvious diagnosis which is common in cases of hyperkeratotic, ulcerative, or vesiculobullous tinea pedis, laboratory diagnosis using superficial skin scrapings^13,14^ or nail clippings^14^ from the infected areas is very helpful. A wet mount of the collected samples is made in potassium hydroxide and observed under the microscope^13,16,45,46^. KOH destroys skin cells leaving fungal cells for examination^46^. Fungal culture methods are the gold standard for the diagnosis of dermatophytosis and are used to confirm the condition due to the similarity of tinea pedis to other diseases^16,47^ and the ineffectiveness of the KOH preparation for severe cases^13^. Severe cases present difficulties with the identification of the fungus in skin scrapings. Misdiagnoses may also occur because of a possible bacterial superinfection. Fungal culture takes one to two weeks for results to come in and is an expensive procedure. The methods are therefore only used when clinical and microscopy diagnosis techniques are inconclusive^46^.

### Complications

Secondary cellulitis, erythema, lymphangitis, maceration, painful vesicles^48^, and fissures are complications that characterize the acute forms of tinea pedis while scaling, peeling, and toe maceration are associated with the chronic forms of the infection^7^. Complications associated with tinea pedis are more frequently associated with other conditions such as diabetes, chronic edema, suppressed immunity, hemiplegia, and paraplegia^49^.

### Prevention

The best way of avoiding any dangers associated with foot ailments and other forms of infections is the maintenance of proper personal hygiene. Patients have to be educated on the importance of foot hygiene to prevent and minimize tinea pedis infections^1^. Effective personal hygiene keeping, dried feet, proper nail care, wearing of well-ventilated shoes with clean-dry socks^1,13^, and sanitizing footwears^50^. One way of ensuring dried shoes is by alternating shoes and socks frequently and using socks made of a material that draws moisture away from the feet. The usage of sandals and flip-flops is also very effective in preventing infection by allowing free movement of air around the feet. Foot powders are recommended for those working or living in areas the fungi have been identified^13^. The disease is prevalent in patients with compromised immunity^51^ such as those infected with Human Immunodeficiency Virus (HIV), alcoholics, and those undergoing organ transplant. Such persons require proper diagnosis as a means of preventing severe tinea pedis. Effective implementation of the preventive measures listed above also helps eradicate infections through interpersonal contacts^1^.

## Antifungal susceptibility testing (AFST)

The ineffectiveness of a drug is a possible cause of resistance to antifungal agents. Two antifungal susceptibility-testing techniques based on broth microdilution methods^52^ are used to determine the minimum inhibitory concentration (MIC) (MIC determination by reference techniques) of drugs used in treating tinea pedis and to detect drug-resistant strains development^19^. Strains with high MIC values are less likely to respond to antifungal agents^52^. AFST is determined by standardized procedures of Clinical and Laboratory Standards Institute (CLSI) and European Committee on Antimicrobial Susceptibility Testing (EUCAST) reference methods^53,54^. The Sensitive Yeast One and E-test methods are used for susceptibility testing.

## Treatment options

Antifungal agents used in the past to manage tinea pedis comprised of inorganic salts such as potassium permanganate and potassium iodide in ointments. Gentian violet, undecanoic, acetylsalicylic acids, and others followed these^19^. The disease, which is mostly on the top layer of the skin, is treated with topical agents and systemic agents when the area of infection is large with several failed attempts^16^, thus, systemic agents are more effective than topical agents^55^. A combination of topical and systemic medication can be more effective. People with underlying health conditions require oral or systemic therapy. The medications used in the treatment of the disease occurs in so many forms such as from creams to shampoo, lacquer, powders, ointments, sprays, and lotions^16^. Topical agents are effective for mild forms of Tinea pedis^1^. Cheaper over-the-counter (OTC) agents work but have socioeconomic implications such as costly. For instance, terbinafine is more effective in the treatment of T. pedis but it is highly costly with one tablet causing about $18.00 and the recommended doses is x times a day. Oral medication is effective for all types of Tinea pedis. There are four classes of systemic and topical antifungal medications used for treating fungal infections and therefore Tinea pedis^56,57^. These are; the polyenes antibiotics, the azole derivatives, the allylamines/thiocarbamates, and the fluoropyrimidines^56,58^. Topical antifungals require about two to four weeks while oral antifungals require about one to two weeks based on the medication chosen. Herbal medications have proved very effective in treating the disease condition^59^, Kakande and his colleagues carried out experimentation using *Tetradenia riparia*^60^.

### The polyenes

These are produced by the *Streptomyces* species and have a broad-spectrum clinical antifungal effect. They work by inhibiting the biosynthesis of ergosterol which is an important component of the fungal plasma membrane by releasing oxidative agents. A typical example is amphotericin B, which may cause such as nephrotoxicity in patients^56^. These are normally given by injection into a vein (intravenous).

### Azoles

They are broad-spectrum synthetic antifungals grouped as imidazole and triazoles based on the number of nitrogen groups. Like polyenes, azoles inhibit the biosynthesis of ergosterol but at the C-14 demethylation step. This inhibitory effect leads to the accumulation of intermediates that inhibits the function of ergosterol in the membrane. This exposes the membrane to damages and its transportability and proliferation are curtailed^56^. Examples of azoles are miconazole, ketoconazole, fluconazole, oxiconazole^61^, econazole, clotrimazole^62^, efinaconazole^63,64^ and luliconazole^65,66^. Azoles are less toxic but some have major or minor side effects. Fluconazole has side effects that are damaging to health^67^. Reported side effects of ketoconazole include reduced production of testosterone and glucocorticoids. They also require about two to four weeks of treatment^14^.

### Clotrimazole

This is an azole with enzyme inhibition property that kills and prevents reinfection of the tinea fungi. Some brands of clotrimazole are Lotrimin and Mycelex^68^. Clotrimazole is applied twice daily for four weeks for effective treatment.

#### Miconazole, ketoconazole, fluconazole and oxiconazole, itraconazole

These antifungals like clotrimazole are azoles with inhibitory effects^69^ on growth enzymes, hence killing dermatophytes and prevents reinfection. The clinically used form of these is Micatin and is applied twice daily for two to four weeks^68^. These have however had the side effect of causing maceration in patients.

Other agents are imidazole, which is effective for interdigital Tinea pedis, and econazole which is effective for cutaneous tinea infections^68^.

#### Allylamines and thiocarbamates

These are synthetic systemic antifungal agents. They inhibit ergosterol biosynthesis by preventing the cyclization of squalene to lanosterol^56^ through squalene epoxidase inhibition^37^. This inhibition leads to a damaging effect on membrane synthesis. They are very effective in treating all forms of Tinea pedis^68^. They are therefore effective in treating dermatophytes^37^. Examples of allylamines are tolnaftate, ciclopirox, butenafine, naftifine, terbinafine with terbinafine the most used clinically.

#### Terbinafine

This is an allylamine that kills fungi and prevents reinfection through growth enzyme inhibition. The common brand name is Lamisil, which exists in spray, and powder forms. It is very effective for treating tinea pedis^37,70^ when applied once daily for a week. Common side effects associated with terbinafine are itching, rubor, and blistering. Fungal resistance to this antifungal is unfortunately established^71^.

#### Tolnaftate

This is an allylamine that inhibits some growth enzymes and prevents infection and reinfection. Tinactin, Desenex spray, Absorbine, and Ting are some common brand names. The cream or gel is applied twice daily for two to six weeks. Some side effects associated with the use of tolnaftate are itching, redness, and peeling^68^.

#### Naftifine (naftin)

They are a broad-spectrum synthetic derivative of allylamine that inhibits sterol biosynthesis for the formation of the fungal cell wall. The application of gel or cream twice daily for 4 weeks is effective for T. rubrum tinea pedis.

#### Butenafine (Mentax)

This is a benzylamine that prevents fungal cell growthgup^72^ by inhibiting sterol synthesis important for cell growth.

Benzyl amine, an allylamine derivative effective for refractory tinea pedis. Ciclopirox inhibits DNA, RNA, and protein synthesis^68^.

#### 5-fluorocytosine (5-FC)

5-FC limited spectrum of activity and is combined with amphotericin B for eliminating candidiasis. Nucleoside triphosphate from 5-FC conversion incorporated into the RNA sequence causes miscoding. It has been used to treat dermatophyte infections including Tinea pedis.

#### Combined steroid/antifungal agents

Combination therapy is sometimes very effective for treating infections and for preventing the development of resistance. A combination of steroids with antifungal agents helps in the treatment of fungal infections. In conditions of no inflammation, the steroid is not needed, hence, monotherapy. A typical example of combination therapy is clotrimazole-betamethasone (Lotrisone)^56^ and miconazole/urea^73^ combinations.

Local ointments and soaps are effective for the treatment of the infection. Some of these ointments are ‘Chocho’ soap and cream, Joy ointment, Whitfield’s ointment, and Castellani’s paint. For effective treatment and avoidance resistance development, treatment must be continuous for at least a week after the clinical clearing of tinea pedis. Other drugs in various developmental stages are echinocandins, pneumocandins, and improved azoles that will target fungal protein and lipid biosynthesis^56^. Other agents used to supplement the antifungals are Aluminum acetate solution, Ammonium lactate lotion, and urea^68^.

### Efficacy of the drugs

The MIC is the mean inhibitory concentration, the mean concentration of the drug at which the organism’s growth is inhibited by 50%. A high MIC value means low potency and vice versa. A comparison of the placebo effect to treatment has shown that treatment is more effective and some tinea pedis left untreated can become severe and lead to complications. Of the drugs used in treating Tinea pedis, terbinafine is the most effective followed by itraconazole though the difference not significant^2^. Fluconazole, griseofulvin, and ketoconazole are very close in potency and to itraconazole. All the drugs have reported side effects with the most common being diarrhea, nausea, headaches, rash, dermatitis, dizziness, and respiratory disorders.

### How to use the medications

To use the topical creams, gels, lacquers, or powders the area of infection and the surrounding parts are cleaned, dried and hands washed to avoid spreading to finger or other body parts^4^. The medicine is applied to the affected area and covered with loose gauze for the free circulation of air. Sandals or flip-flops must be worn after application or if possible patients should walk barefooted after medication. Failures of treatment should be reported to a medical practitioner or dermatologist six weeks after treatment.

## Resistance

Fungal resistance occurs when the fungus can survive in the presence of the drug made against it or appropriate therapy. It occurs as a result of the alteration of one or more genes making a previously susceptible population resistant^58^. Resistance commonly develops from incomplete medication regimens or the misuse of antifungal medications^19,74^. Treatment of patients with underlying health conditions such as AIDS and diabetes^17^ and immunocompromised patients such as those undergoing cancer therapy and transplantation^56^ is complicated and also leads to drug resistance. Besides, resistance may develop but remain undetected due to low routine susceptibility testing hence complicating the situation. Even though fungal drug resistance is low compared to bacteria, the current rise in resistance poses a serious threat to humanity, animals, plants, and the ecosystem^67^. Margaret Hudson (2001) reported a yearly record of 2.8 million antibiotic-resistant infections and 35,000 deaths in the United States. Resistance to allylamines^37^ and azoles (*Candida albicans* to fluconazole) have been reported^58^ and linked to the overuse of terbinafine. Some antifungals are more vulnerable to resistance than others. TruMDR1 and TruMDR2-ATP-binding genetic cassettes are linked to resistance development and pathogenic effects of dermatophytes^37^.

## Mechanism of drug resistance

Resistance mechanisms are the various defense strategies employed by microbes to overcome and survive^19^ the host defenses, antibiotics, and other chemotherapies. The invading germ develops strategies such as alteration of surface lipids or proteins, use of different synthetic routes or synthesize compounds that counter the effect of the host’s defense of antibiotics. Some mechanisms employed in resistance development include metabolic degradation of drugs, mutations, recombination, alteration of the drug target site, and target molecule overexpression^37^. Resistance development reduces the potency of drugs.

### Polyenes

Polyene resistance is from the alteration of membrane lipids and enhanced catalase activity, which helps reduce oxidative stress on the fungal cell. Amphotericin B has known secondary clinical resistance by some fungal strains (*Candida sp*.) even though intrinsic clinical resistance is not common. Polyenes are very toxic to the host^75^.

### Azoles

Resistance to azoles is very common, particularly in immunocompromised patients. Resistance develops from the alteration of membrane sterols leading to reduced membrane permeability. Mechanisms involved are stress adaptation, increased drug efflux, and overexpression of essential enzymes^19^. Resistance has been reported in C. neoformans^56^ and clinical Trichophyton spp^76^.

### Allylamines

Resistance to allylamines develops through mutation of enzymes required in the action of antifungals, stress adaptation, increased drug efflux, and drug degradation^19^. Terbinafine resistance in T. rubrum was clinically confirmed by Colin et al. and Ditte et al. to be a result of amino acid substitution in squalene epoxidase^77,78,79^.

### 5-FC

Occurs as a result of cytosine deaminase inhibition or mutations of enzymes required for 5-FC functioning^56^.

## Methods of controlling resistance

Infection prevention and control, tracking and data sharing, good and easily accessible antifungals, vaccines, and maintenance of personal and environmental hygiene^67^ prevent resistance development. Also, resistance development can be controlled by delaying systemic azole therapy, effective dosing of medications, use of combination therapy with different actions, and use of systemic azoles with stimulators (e.g. cytokines) and efflux pump inhibitors^58^. Patient education to religiously follow prescribed treatment regimens also helps to control drug resistance.

## New drug synthesis

In new drug synthesis, of necessary consideration is the various mechanisms used in resistance development. Potent drugs should have no or less effect on the host. New drugs should be able to accumulate and persist at the site of application for a longer time^19^.

In conclusion, there is a need to using combination therapy as well as herbal treatment that has been shown to exert synergistic effectiveness.

## Data Availability

N/A

